# EEG-Based Prediction of rTMS Treatment Response in Depression: Nonlinear Features and Machine Learning with Minimal Electrode

**DOI:** 10.1101/2025.10.30.25339032

**Authors:** Chun-Hung Chang, Alexander T. Sack, Che-Sheng Chu, Hsin-An Chang

## Abstract

**Background:** Repetitive transcranial magnetic stimulation (rTMS) is an established intervention for treatment-resistant depression, but response rates remain highly variable and reliable predictors of outcome are lacking. Resting-state electroencephalography (EEG), combined with machine learning, represents a cost-effective and accessible approach for individualized treatment planning.

**Methods:** One hundred patients with major depressive disorder (MDD; 51 responders, 49 non-responders) underwent pre-treatment resting-state EEG prior to rTMS. Recordings were obtained using either 8 or 30 electrodes. Features including band power, coherence, phase-lag index (PLI), and phase-locking value (PLV) were extracted. Machine learning models were constructed using two-stage feature selection (recursive feature elimination with support vector classifier and sequential backward selection) and linear discriminant analysis (LDA). Model performance was assessed with 5-fold cross-validation and 1,000 shuffle iterations.

**Results:** Multi-feature EEG models consistently outperformed single-feature approaches in predicting rTMS treatment response. The best-performing model, based on only 8 electrodes, achieved an accuracy of 78.9% and an AUC of 73.3%, surpassing the 30-electrode configuration. PLI emerged as the strongest individual predictor, but combining multiple EEG features substantially improved classification. LDA provided the most stable performance across limited datasets.

**Conclusions:** Resting-state EEG combined with machine learning can reliably predict rTMS treatment response in MDD. Notably, accurate prediction was achieved with a minimal 8-electrode montage, supporting the clinical feasibility of low-cost EEG assessments. This approach may facilitate personalized treatment selection and improve rTMS outcomes in routine psychiatric care.

## Introduction

Major Depressive Disorder (MDD) is a debilitating psychiatric condition with a steadily increasing global prevalence. According to the World Health Organization, MDD was the third leading cause of disease burden worldwide in 2008 and is projected to become the leading cause by 2030(Malhi and Mann, 2018). While pharmacological treatments, particularly antidepressants, remain the primary approach for managing MDD, approximately one-third of patients fail to achieve remission despite receiving adequate antidepressant therapy(Gaynes et al., 2008). Moreover, approximately one-third of patients discontinue antidepressant treatment due to intolerable side effects. In a large retrospective cohort study involving 7,723 individuals, 36.9% stopped at least one antidepressant for this reason(Garcia-Marin et al., 2023). These challenges highlight the urgent need for alternative, effective treatment strategies and predictive tools that can guide personalized interventions for depression.

Repetitive transcranial magnetic stimulation (rTMS) has emerged as an innovative, non-invasive, and non-pharmacological treatment for patients with treatment-resistant depression(Hallett, 2007). Approved by the U.S. Food and Drug Administration (FDA), rTMS is now widely adopted in clinical settings (Lefaucheur et al., 2020). However, despite its overall therapeutic benefits, the response rate of rTMS remains approximately 50%(Kennedy et al., 2016; Brunoni et al., 2017), indicating a large variability between patients with regard to treatment success. Moreover, there remains a lack of consensus on how to predict individual treatment outcomes. To address this gap, various neurobiological and physiological biomarkers have been explored to enhance precision medicine approaches. Modalities such as functional magnetic resonance imaging (fMRI), genetic markers, near-infrared spectroscopy (NIRS), and electroencephalography (EEG) have shown promise in identifying responders to rTMS(Klooster et al., 2024).

Among the available biomarker modalities, EEG offers a particularly attractive option due to its non-invasive nature, relatively low cost, high temporal resolution, and ease of implementation in clinical environments. As a measure of brain electrical activity, EEG can reflect functional abnormalities and network dynamics associated with depression(Fingelkurts and Fingelkurts, 2015). Recent studies have begun to leverage EEG features—such as band power, coherence, and phase-based metrics—to predict treatment outcomes in depression, including responses to rTMS(Watts et al., 2022; Corlier et al., 2019; Bailey et al., 2019; Erguzel et al., 2015; Bailey et al., 2018; Hasanzadeh et al., 2019). One study involving 55 MDD patients employed an artificial neural network (ANN) and reported an overall classification accuracy of 89.09%(Erguzel et al., 2015). Another study applied the k-nearest neighbors (KNN) algorithm to a sample of 46 MDD patients and achieved an accuracy of 91.3%(Hasanzadeh et al., 2019). Furthermore, a recent meta-analytic review incorporating seven rTMS studies reported that the mean classification accuracy for predicting rTMS treatment response ranged from 68.5% to 91.3%, with a pooled accuracy of 85.70% (95% CI: 77.45–94.83) (Watts et al., 2022). However, the optimal electrode configuration and feature combination for predictive modeling remain unclear. This study addresses these gaps by comparing the predictive performance of EEG models based on 8-electrode and 30-electrode configurations and evaluating both multi-feature and single-feature analysis approaches.

In this study, we aimed to determine whether a reduced set of EEG electrodes could effectively predict rTMS treatment response using an optimized AI-based classification model. By applying a two-stage feature selection process and linear discriminant analysis (LDA), we investigated whether simplified EEG configurations, when combined with nonlinear signal features, could match or exceed the predictive accuracy of full-cap setups.

## Methods

### Participants

The single-center prospective cohort study was conducted at Tri-Service General Hospital (TSGH) and the patients were recruited between March 18, 2024 and January 20, 2025. All patients referred for clinical rTMS at the TSGH Brain Stimulation Unit were consecutively approached for potential study participation. Inclusion criteria were being 18-65 years, having DSM-5 (the Diagnostic and Statistical Manual of Mental Disorders, Fifth Edition)-defined major depressive disorder (MDD), meeting the criteria for treatment-resistant depression (TRD) defined as no clinical response to at least two adequate trials of a major class of antidepressants, and having provided written informed consent. Exclusion criteria were patients deemed unsuitable for participating in this study by the medical judgment of the investigators due to pregnancy or other reasons (e.g., lack of competence to consent to research or any other contraindications to receiving rTMS). The Maudsley Staging Method (MSM) was used to define and stage treatment resistance for each participant with MDD. The study was approved by the local research ethics committees (IRB number: B202405005) and was conducted in accordance with the latest revision of the Declaration of Helsinki. Written informed consent was obtained from all subjects.

### Accelerated piTBS

The accelerated piTBS sessions were delivered using the Magstim Rapid2 stimulator (Magstim Company, Ltd, Whitland, Wales, UK, SA340HR) with a 70-mm air-cooled figure-eight coil (Magstim D70 Air film coil). The protocol consists of 3-pulse 50-Hz bursts given every 200 ms (at 5 Hz) for 2 s at 8-s intervals for 60 cycles. A 2-s train of stimulation was repeated every 10 s for a total of 1800 pulses per session. The intensity of stimulation was set at 90% resting motor threshold (RMT), as measured from the right first dorsal interosseous muscle by a handheld 700-mm figure-of-eight coil (Magstim D70 alpha coil). The Beam F3 method was used for coil positioning to target the left dorsolateral prefrontal cortex. The accelerated piTBS was scheduled for 3 sessions per working day for 2 weeks (i.e., a total of 30 sessions over 2 weeks). Three sessions of the piTBS per day were separated by 60 mins.

### Outcome measures

Hamilton depression rating scale (HAMD) was used to measure clinician-rated depression severity. At the end of stimulation, participants were divided into HAMD score responders and non-responders. A HAMD responder was defined as a subject who had a ≥50% reduction from baseline in HAMD total score. The self-reported overall severity of depression and anxiety was assessed using the Patient Health Questionnaire (PHQ-9)(Chung et al., 2023), and Generalized Anxiety Disorder 7 (GAD-7)(Toussaint et al., 2020), respectively. Subjective cognitive dysfunction in MDD patients was measured with the Taiwan Cognition Questionnaire (TCQ) (Yen et al., 2022).

### Modeling and Classification Performance

In this study, we developed predictive models using both 8-channel and 30-channel EEG configurations. Model performance was evaluated using 5-fold cross-validation, and to ensure stability and reliability of the results, each experiment was repeated with 1,000 random shuffles.

The 8-electrode configuration was based on the design of the SEA Index, an AI-powered EEG-based stress assessment system developed by HippoScreen Neurotech Corp., which has received regulatory approval from the Taiwan Food and Drug Administration (TFDA) as a Software as a Medical Device (SaMD) for aiding in the assessment of suspected depression.

The selected electrodes—CP3, T5, FC3, FT8, F3, Cz, F8, and Fz—matched those used in the TFDA-approved system and were used for subsequent analysis(Wu et al., 2021).

### EEG Acquisition

Visual stimuli were presented on a 24-inch flat-panel monitor, with participants seated approximately 70 cm from the screen. EEG signals were recorded using an 8-channel Waveguard cap connected to an HNC amplifier (provided by HippoScreen Neurotech Corp.). The amplifier gain was set to 50×, with a 24-bit A/D resolution and a sampling rate of 500 Hz.

Electrodes were composed of aluminum/chloride silver (Al/AgCl) and positioned according to the international 10–20 system. Conductive gel (Gelaid, Nihon Kohden) was applied to enhance signal conductivity. The reference electrode was placed on the right mastoid (A2 position).

### Data Collection and Preprocessing

For each participant, a 90-second resting-state EEG recording was obtained during an eyes-open, relaxed condition. A timer cue was used to initiate the session, during which a black fixation cross on a gray background appeared on the screen. Participants were instructed to maintain relaxed gaze on the cross throughout the 90-second interval.

During preprocessing, the EEG signals were re-referenced to the Cz electrode and digitally filtered using a bandpass filter ranging from 2 to 60 Hz. The continuous EEG recordings from seven electrodes were then segmented into fifteen non-overlapping epochs, each lasting 6 seconds, for subsequent analysis.

### Dataset Preparation and AI Model Training

In this study, we developed an artificial intelligence (AI) model to predict the therapeutic response to repetitive transcranial magnetic stimulation (rTMS) using EEG data. Of the initial 100 participants, EEG recordings from 6 individuals were excluded due to poor signal quality, resulting in a final dataset of 94 valid cases. The data were randomly divided into training and testing sets in an 8:2 ratio, yielding 75 participants in the training set and 19 in the testing set.

To ensure comparability between responders and non-responders within both sets, statistical tests were conducted on age and sex distributions. In the training dataset, no significant differences were found in sex (p = 0.3169, Cohen’s d = 0.2877) or age (p = 0.4838, d = 0.2193) between the two groups. Similarly, in the testing dataset, sex (p = 1.0000, d = 0.2323) and age (p = 0.3529, d = 0.4388) distributions were also statistically comparable. These results indicate that the demographic characteristics of the training and testing sets were balanced, minimizing potential confounding effects on model performance.

### EEG Feature Extraction

The EEG features used in this study included the following: Band Power (BP)

Relative Band Power (RP)

Ratio Band Power

Coherence (Coh)

Phase-Lag Index (PLI)

Phase-Locking Value (PLV)

### Band Power (BP)

Power spectral density (PSD) was computed using the Fast Fourier Transform (FFT). Absolute band power was calculated for five standard frequency bands: delta (2–4 Hz), theta (4–8 Hz), alpha (8–13 Hz), beta (13–30 Hz), and gamma (30–45 Hz). To account for the wide magnitude range of the computed BP values, the mean BP was log-transformed to obtain log-transformed band power (log BP) for further analysis.

### Relative Band Power (RP)

Relative band power is a feature that quantifies the proportion of energy within a specific frequency band relative to the total EEG signal power. It is commonly used to evaluate the contribution of individual frequency components across different brain regions.

Three types of RP were analyzed:

RP Type I (Power Difference Ratio):

This method calculates the difference in power between two electrode sites relative to their total power, emphasizing the contrast in activity between the two regions. The formula is:

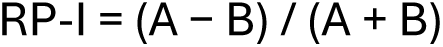

where A and B represent the band power values at two different electrode sites. This ratio highlights asymmetry and energy contrast between paired channels.

RP Type II (Power Ratio):

This measure expresses the ratio of power between two electrode sites, providing a direct comparison of their relative energy contributions. The formula is:

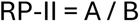

This approach captures the proportional distribution of signal power between regions.

RP Type III (Logarithmic Power Difference):

To reduce the influence of extreme power values and better capture relative differences on a logarithmic scale, the third variant computes the difference between log-transformed power values:

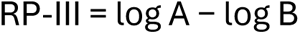

This method offers improved stability and sensitivity, particularly when power differences are large.

### Ratio Power (RP)

Ratio power is used to assess the relative contribution of specific EEG frequency bands to overall brain activity. In this study, ratio power was calculated for five standard frequency bands: delta (2–4 Hz), theta (4–8 Hz), alpha (8–13 Hz), beta (13–30 Hz), and gamma (30–45 Hz). The total power was computed across the full spectrum from 2 to 50 Hz.

The ratio power for each band was computed as the proportion of band-specific power to the total power within the 2–50 Hz range, using the following formula:

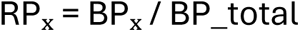

where RP_x_ denotes the ratio power of frequency band X, BP_x_ is the absolute band power of frequency band X, and BP_total is the sum of power across the 2–50 Hz range.

This approach allows for a normalized comparison of activity across frequency bands, contextualized within the overall spectral power of the EEG signal.

### Coherence

EEG coherence was calculated using the Fast Fourier Transform (FFT) to evaluate the linear relationship between signals from pairs of electrodes within specific frequency bands. For each electrode pair (i, j) and frequency band f, the squared coherence was computed based on the following equation:

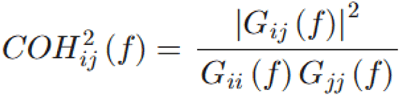

where *COH_ij_* (*f*)denotes the coherence at frequency *f*, *G_ij_*(*f*)represents the averaged cross-power spectral density between electrodes *i* and *j*, and *G_ii_*(*f*) and *G_jj_*(*f*) denote the averaged auto-power spectral densities of electrodes *i* and *j*, respectively.

Coherence values range from 0 to 1, with higher values indicating stronger linear synchronization between the EEG signals of the two electrodes.

### Phase Lag Index (PLI)

The Phase Lag Index (PLI) is a metric used to quantify phase synchronization between two EEG signals by measuring the consistency of phase differences over time. Rather than assessing amplitude correlations, PLI focuses on the temporal stability of phase lead or lag between signal pairs.

PLI values range from 0 to 1, where a value of 0 indicates no consistent phase relationship (i.e., random phase differences), and a value of 1 indicates perfect, time-consistent phase locking between the signals. Higher PLI values suggest stronger functional connectivity based on phase relationships, reflecting neural coupling with minimal influence from volume conduction or common sources.

Phase-Locking Value (PLV):

The Phase-Locking Value (PLV) is a measure used to assess the degree of phase synchronization between two EEG signals, focusing on the consistency of their phase relationship over time. PLV evaluates how stable the phase difference is within a defined time window. A PLV value close to 1 indicates highly consistent phase alignment between signals, suggesting strong functional connectivity. Conversely, a value near 0 reflects high variability in phase difference, indicating weak or no synchronization.

### Feature Selection Strategy

To enhance model performance and reduce dimensionality, a two-stage feature selection approach was employed:

### Recursive Feature Elimination with Support Vector Classifier (RFE-SVC)

RFE is a backward feature elimination technique that recursively trains a model and removes the least important features based on their contribution to classification performance. This process continues until the optimal subset of features is identified.

### Sequential Backward Selection (SBS) with Linear Discriminant Analysis (LDA)

In the second stage, SBS(Saeys et al., 2007; Tang et al., 2022) was applied using an LDA(Ricciardi et al., 2020) classifier to further refine the selected features. Considering the small sample size, the number of retained features was set to (N − 1), where N is the number of training samples. SBS begins with the full feature set and sequentially removes features that contribute the least to model performance, ultimately retaining those with the highest classification utility.

This two-tiered selection process was designed to maximize model accuracy while ensuring the robustness and interpretability of the selected EEG features.

## Results

The cohort has 100 TRD patients who received 30 sessions of accelerated piTBS. The participants’ demographic and clinical baseline data are presented in Table 1. The mean of the MSM total score was 10.06 ± 1.13. At the end of stimulation, 51 % of the study participants were HAMD responders (n = 51). Treatment resistance for responders and non-responders before stimulation was not significantly different (MSM total score: 9.88 ± 0.84 versus 10.24 ± 1.35, p = 0.11).

**Table 1.** Demographic and clinical information.

|  | **Non-responders**<br>**n=49** | **Responders**<br>**n=51** | **p-Values** | **Effect Size** |
| --- | --- | --- | --- | --- |
| **Gender** | 10 F, 39 M | 17 F, 34 M | 0.2187 | 0.2913 |
| **Age** | 43 (±13.69) | 46.57 (±12.73) | 0.2156 | 0.2702 |
| **Pre HAMD** | 26.53 (±5.99) | 25.11 (±5.55) | 0.2312 | 0.2446 |
| **Post HAMD** | 19.44 (±6.69) | 9.45 (±2.88) | 7.55E-15 | 1.9521 |
| **Pre PHQ-9** | 20 (±3.42) | 19.78 (±3.38) | 0.7316 | 0.0633 |
| **Post PHQ-9** | 14.16 (±4.09) | 7.49 (±1.46) | 1.45E-16 | 2.1857 |
| **Pre TCQ** | 9.12 (±2.42) | 9.03 (±2.81) | 0.9805 | 0.0317 |
| **Post TCQ** | 8.53 (±2.33) | 6.66 (±2.04) | 4.90E-05 | 0.8497 |
| **Pre GAD-7** | 12.51 (±3.75) | 12.76 (±2.49) | 0.7444 | 0.0802 |
| **Post GAD-7** | 7.91 (±2.54) | 4.92 (±1.65) | 1.09E-08 | 1.3995 |

aChi-square test for the categorical data; Student's t-test for continuous variables.
Note: a) Chi-square test; b) ANOVA test; c) Mann–Whitney U test.

### Performance of the AI Model Using 8-Electrode EEG Configuration

When using the 8-electrode configuration, the best performance during 5-fold cross-validation was achieved with 27 selected features. The model yielded a training accuracy of 95.89%, with a sensitivity of 97.50% and specificity of 94.29%. After averaging across 1,000 shuffles, the training set achieved a mean accuracy of 86.36%, sensitivity of 87.25%, and specificity of 85.48%.

For the independent test set, the model achieved a classification accuracy of 78.89%, with a sensitivity of 80.00% and specificity of 77.78%. The area under the ROC curve (AUC) for the test set was 73.33%, with a positive predictive value (PPV) of 80.00% and a negative predictive value (NPV) of 77.78% (Figure 2).

**Figure 1.**
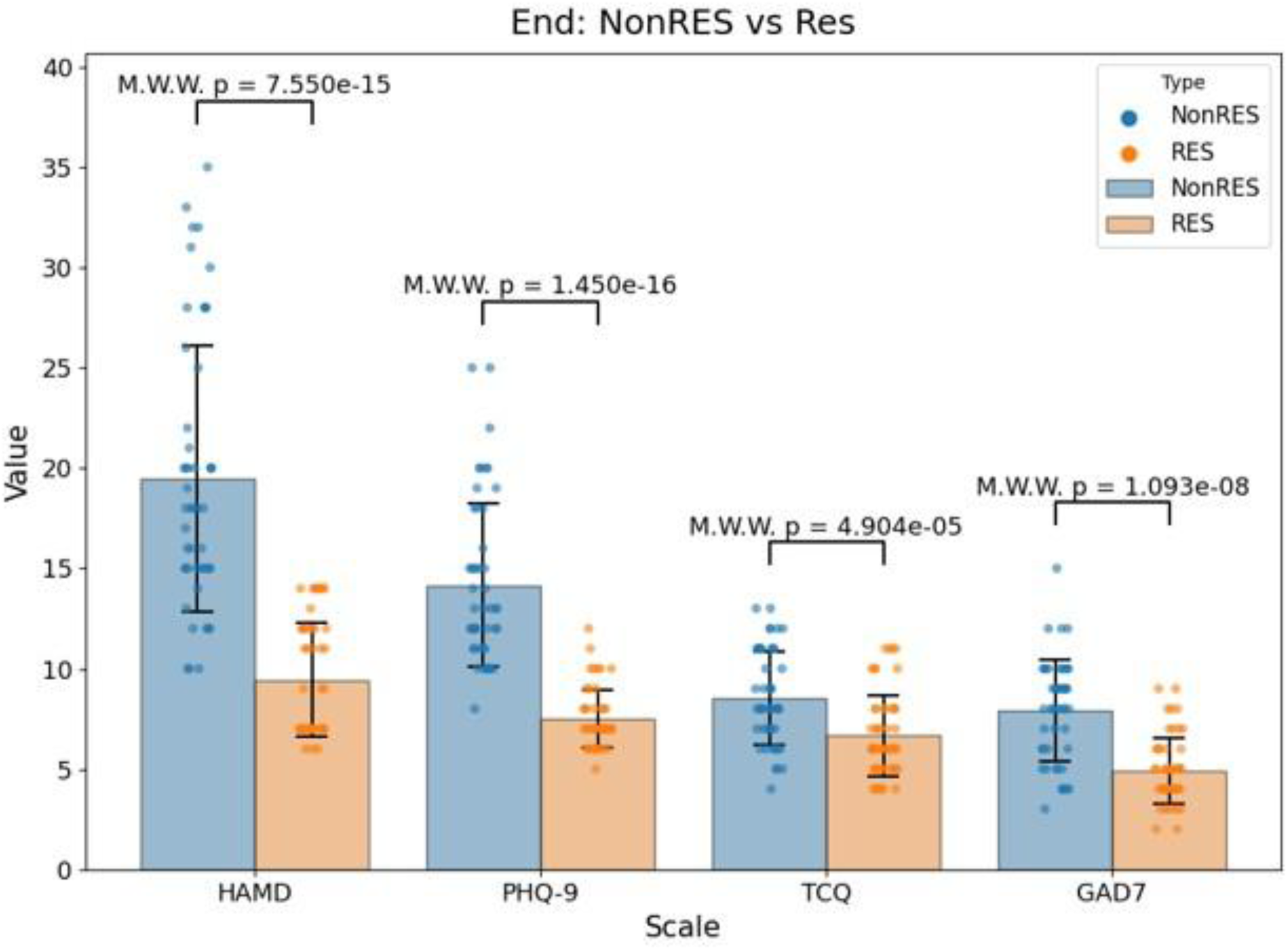
Group Differences in Post-Treatment Clinical Scores Between Responders and Non-Responders to rTMS in Depression. Note: Bar plots illustrate the distributions of HAMD, PHQ-9, TCQ, and GAD-7 scores at the end of rTMS treatment. Responders (RES) showed significantly lower scores across all scales compared to non-responders (NonRES), indicating greater symptomatic improvement. Mann–Whitney–Wilcoxon (M.W.W.) test p-values are shown for each comparison.

**Figure 2.**
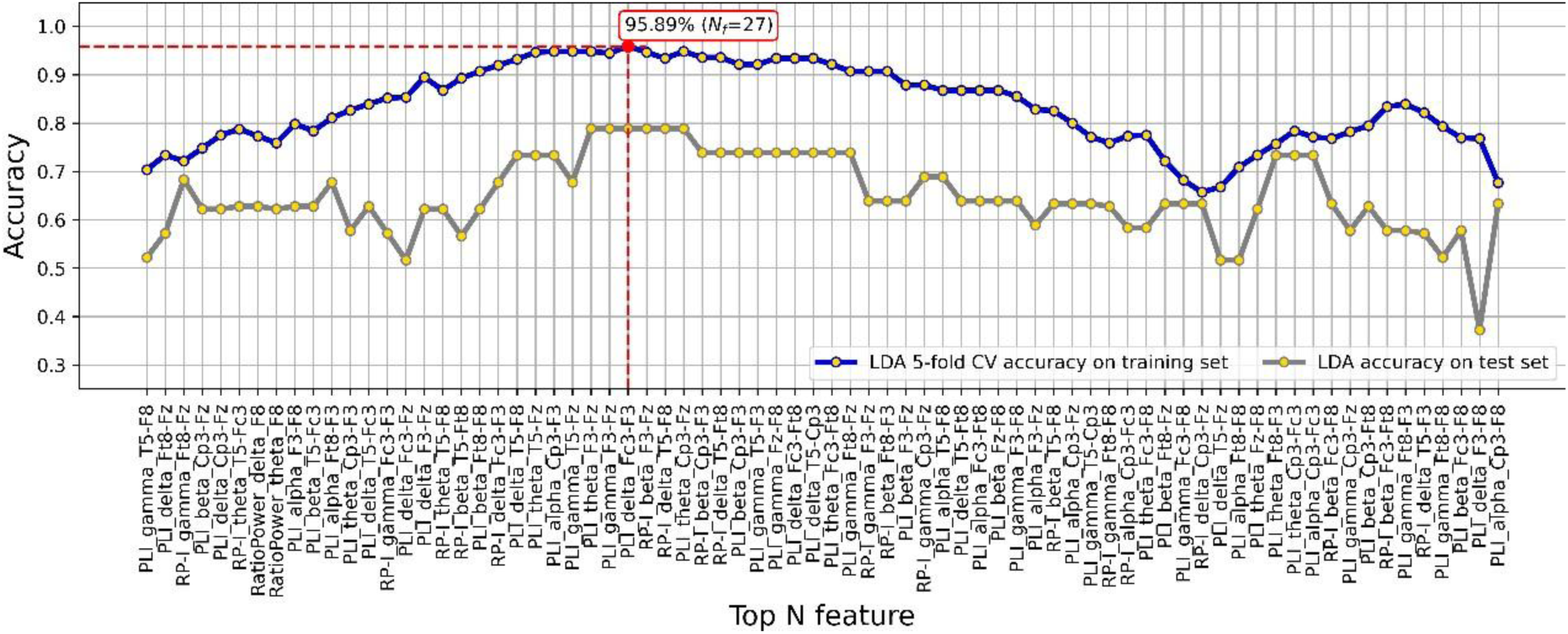
Classification accuracy across top-N EEG features using 8-channel configuration with RFE-SVC and SBS feature selection. The blue curve represents 5-fold cross-validation (CV) accuracy on the training set using a Linear Discriminant Analysis (LDA) classifier, while the yellow curve indicates classification accuracy on the independent test set. The highest training accuracy (95.89%) was achieved using the top 27 features. This feature subset was identified through a two–stage selection process involving RFE-SVC and SBS. Feature labels on the x–axis represent specific EEG metrics and electrode pairs contributing to model performance.

### Performance of the AI Model Using 30-Electrode EEG Configuration

When modeling with the 30-electrode EEG configuration, the optimal performance was achieved using 26 selected features. Under 5-fold cross-validation, the model attained perfect classification performance on the training set, with 100.00% accuracy, sensitivity, and specificity.

To assess the robustness of this result, the model was further evaluated using 1,000 randomized shuffles. The average training accuracy was 94.98%, with a sensitivity of 94.86% and specificity of 95.09%. On the independent test set, the model achieved an accuracy of 73.33%, sensitivity of 80.00%, and specificity of 66.67%. The area under the receiver operating characteristic curve (AUC) for the test set was 73.33%, with a positive predictive value (PPV) of 72.72% and a negative predictive value (NPV) of 75.00% (Figure 3).

**Figure 3.**
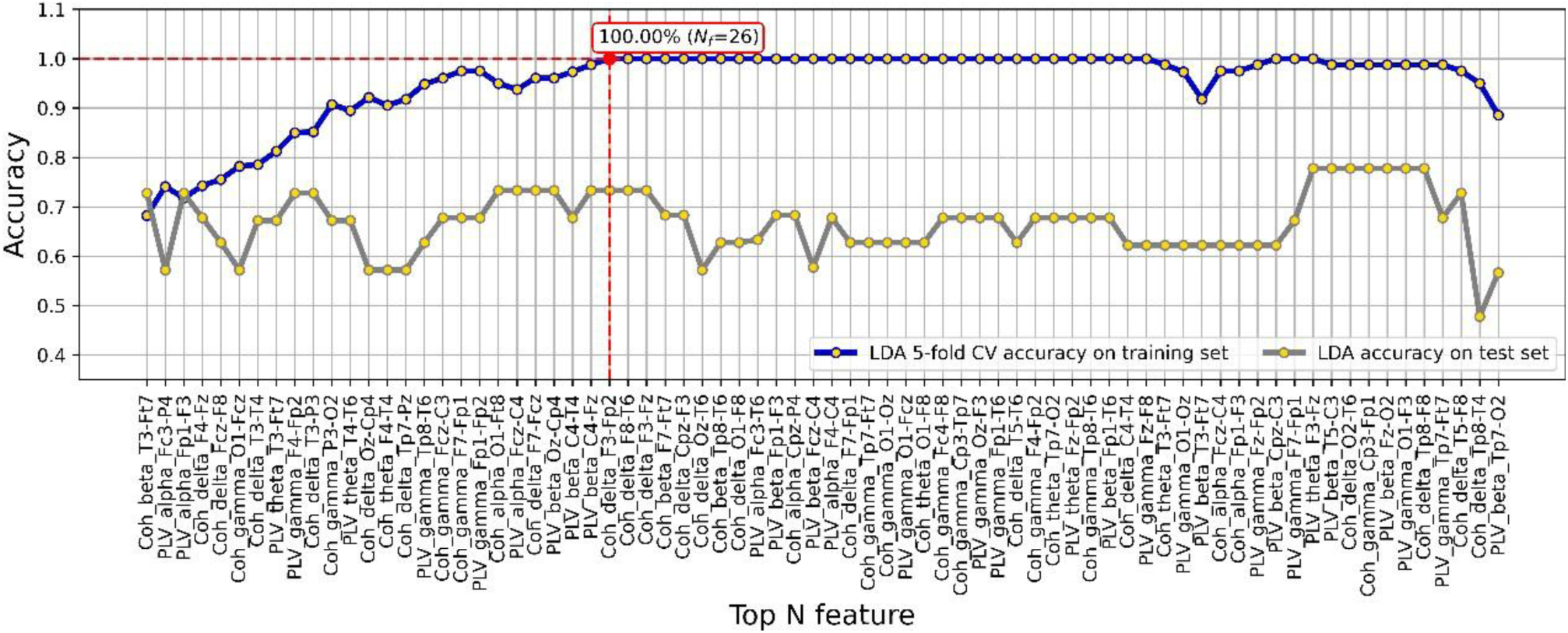
Classification accuracy across top-N EEG features using 30-electrode configuration with RFE-SVC and SBS feature selection. The blue curve shows 5-fold cross-validation (CV) accuracy on the training set using a Linear Discriminant Analysis (LDA) classifier, while the yellow curve represents classification accuracy on the independent test set. The model achieved perfect classification on the training set (100.00%) using the top 26 features. Feature labels on the x-axis denote specific EEG connectivity metrics (Coherence and PLV) and corresponding electrode pairs contributing to model performance.

### Comparative Performance of 8-Channel and 30-Channel EEG Models

The results indicate that although the 30-electrode model outperformed the 8-electrode model on the training set, their performance on the test set was largely comparable. Specifically, the 8-electrode model achieved an area under the curve (AUC) of 73.33%, with a positive predictive value (PPV) of 80.00% and a negative predictive value (NPV) of 77.78%. In contrast, the 30-electrode model yielded the same AUC (73.33%) but slightly lower predictive values (PPV = 72.72%, NPV = 75.00%). These findings suggest that while increasing the number of electrodes may enhance model fitting during training, it does not necessarily lead to improved generalization performance on unseen data.

### Single-Feature Classification Performance

To assess the individual predictive power of each feature, Linear Discriminant Analysis (LDA) classifiers were applied to compute classification metrics across both the training and test datasets. The classification performance of each feature was evaluated under both the 8-electrode and 30-electrode configurations. The results, summarized in the corresponding table, include classification rate (CR), true positive rate (TPR, sensitivity), true negative rate (TNR, specificity), and area under the curve (AUC) for each feature.

### Classification Performance of Individual Features under 8-Channel and 30-Channel Configurations

Under the 8-channel configuration, the Phase Lag Index (PLI) feature demonstrated the best classification performance in the training set, achieving a classification rate (CR) of 87.86%, sensitivity (TPR) of 87.14%, and specificity (TNR) of 88.57%. In the test set, PLI also performed well, with a CR of 73.33%, TPR of 80.00%, and TNR of 66.67% (Table 3).

**Table 2.** Demographic characteristics of responders and non-responders in the training and testing datasets. No significant differences in gender or age were observed between groups, supporting balanced distribution for AI model development.

| Variable (Train n=75) | Res, (n=39) | NonRes, (n=36) | p-Values | Effect Size |
| --- | --- | --- | --- | --- |
| Gender | 15 F, 24 M | 9 F, 27 M | 0.3169 | 0.2877 |
| Age | 46.62 (±12.03) | 43.72 (±14.00) | 0.4838 | 0.2193 |

| Variable (Test n=19) | Res, (n=10) | NonRes, (n=9) | p-Values | Effect Size |
| --- | --- | --- | --- | --- |
| Gender | 2 F, 8 M | 1 F, 8 M | 1.0000 | 0.2323 |
| Age | 44.8 (±14.86) | 38.33 (±12.83) | 0.3529 | 0.4388 |

**Table 3.** Classification Performance of Individual EEG Features Using LDA Classifier in 8-Channel and 30-Channel Configurations.

| Feature | Train |  |  | Test |  |  |  |
| --- | --- | --- | --- | --- | --- | --- | --- |
|  | CR | TPR | TNR | CR | TPR | TNR | AUC |
| **(a) 8 CH** |  |  |  |  |  |  |  |
| Coh | 86.61% | 89.64% | 83.57% | 52.78% | 50.00% | 55.56% | 51.00% |
| LogBP | 70.54% | 76.79% | 64.29% | 56.67% | 80.00% | 33.33% | 69.00% |
| PLI | 87.86% | 87.14% | 88.57% | 73.33% | 80.00% | 66.67% | 68.00% |
| PLV | 74.82% | 80.00% | 69.64% | 57.22% | 70.00% | 44.44% | 68.00% |
| RatioPower | 71.96% | 71.43% | 72.50% | 36.67% | 40.00% | 33.33% | 42.00% |
| RP-I | 81.07% | 81.79% | 80.36% | 62.22% | 80.00% | 44.44% | 76.00% |
| RP-II | 81.25% | 84.64% | 77.86% | 46.67% | 60.00% | 33.33% | 39.00% |
| RP-III | 70.18% | 74.29% | 66.07% | 67.78% | 80.00% | 55.56% | 59.00% |
| **(b) 30 CH** |  |  |  |  |  |  |  |
| Coh | 96.07% | 95.00% | 97.14% | 63.89% | 50.00% | 77.78% | 79.00% |
| LogBP | 92.50% | 85.00% | 100.00% | 57.78% | 60.00% | 55.56% | 64.00% |
| PLI | 100.00% | 100.00% | 100.00% | 73.33% | 80.00% | 66.67% | 78.00% |
| PLV | 100.00% | 100.00% | 100.00% | 62.22% | 80.00% | 44.44% | 71.00% |
| RatioPower | 80.00% | 81.79% | 78.21% | 43.33% | 20.00% | 66.67% | 38.00% |
| RP-I | 100.00% | 100.00% | 100.00% | 68.89% | 60.00% | 77.78% | 69.00% |
| RP-II | 100.00% | 100.00% | 100.00% | 68.33% | 70.00% | 66.67% | 61.00% |
| RP-III | 96.25% | 95.00% | 97.50% | 57.78% | 60.00% | 55.56% | 56.00% |

Classification rate (CR), true positive rate (TPR), true negative rate (TNR), and area under the curve (AUC) are reported for each feature on both training and test datasets. Results are shown separately for 8-channel (a) and 30-channel (b) EEG configurations.

**Table 4.** Comparison of Six Classifiers in Predicting rTMS Treatment Outcomes: Performance Across 8-Channel and 30-Channel EEG Configurations.

| Model | Train |  |  | Train (Shuffle 1000 times) |  |  | Test |  |  |  |  |  |
| --- | --- | --- | --- | --- | --- | --- | --- | --- | --- | --- | --- | --- |
|  | CR | TPR | TNR | CR | TPR | TNR | CR | TPR | TNR | AUC | PPV | NPV |
| **(a) 8 CH** |  |  |  |  |  |  |  |  |  |  |  |  |
| K-Nearest Neighbors | 74.64% | 77.14% | 72.14% | 70.30% ± 3.37% | 55.95% ± 4.94% | 84.65% ± 4.31% | 72.78% | 90.00% | 55.56% | 72.22% | 69.23% | 83.33% |
| Linear Discriminant Analysis | 95.89% | 97.50% | 94.29% | 86.36% ± 3.26% | 87.25% ± 3.94% | 85.48% ± 4.72% | 78.89% | 80.00% | 77.78% | 73.33% | 80.00% | 77.78% |
| Naive Bayes | 80.00% | 76.79% | 83.21% | 76.80% ± 3.22% | 85.60% ± 3.93% | 68.01% ± 4.69% | 73.33% | 80.00% | 66.67% | 76.67% | 72.72% | 75.00% |
| Logistic Regression | 78.39% | 78.93% | 77.86% | 69.75% ± 2.78% | 82.73% ± 4.47% | 56.78% ± 3.15% | 73.89% | 70.00% | 77.78% | 72.00% | 77.78% | 70.00% |
| Support Vector Machine | 80.54% | 80.00% | 81.07% | 72.98% ± 3.86% | 75.49% ± 5.63% | 70.48% ± 4.94% | 78.33% | 90.00% | 66.67% | 74.44% | 75.00% | 85.71% |
| eXtreme Gradient Boosting | 84.11% | 90.00% | 78.21% | 63.79% ± 4.61% | 69.33% ± 6.53% | 58.25% ± 6.84% | 72.78% | 90.00% | 55.56% | 73.23% | 69.23% | 83.33% |
| **(b) 30 CH** |  |  |  |  |  |  |  |  |  |  |  |  |
| K-Nearest Neighbors | 80.00% | 82.14% | 77.86% | 67.73% ± 3.84% | 69.41% ± 5.29% | 66.04% ± 5.08% | 74.44% | 60.00% | 88.89% | 79.00% | 85.71% | 66.67% |
| Linear Discriminant Analysis | 100.00% | 100.00% | 100.00% | 94.98% ± 2.30% | 94.86% ± 2.67% | 95.09% ± 3.32% | 73.33% | 80.00% | 66.67% | 73.33% | 72.72% | 75.00% |
| Naive Bayes | 88.93% | 95.00% | 82.86% | 87.70% ± 2.10% | 88.50% ± 2.80% | 86.91% ± 2.94% | 73.89% | 70.00% | 77.78% | 78.89% | 77.78% | 70.00% |
| Logistic Regression | 86.79% | 84.64% | 88.93% | 92.15% ± 2.59% | 95.99% ± 2.45% | 88.31% ± 4.25% | 78.89% | 80.00% | 77.78% | 75.56% | 80.00% | 77.78% |
| Support Vector Machine | 94.82% | 95.00% | 94.64% | 90.23% ± 2.99% | 91.20% ± 3.53% | 89.26% ± 4.58% | 83.89% | 90.00% | 77.78% | 86.67% | 81.81% | 87.50% |
| eXtreme Gradient Boosting | 82.50% | 87.14% | 77.86% | 69.57% ± 4.11% | 72.46% ± 5.43% | 66.68% ± 6.23% | 73.33% | 80.00% | 66.67% | 73.33% | 72.73% | 75.00% |

The performance metrics presented in this table include classification rate (CR), true positive rate (TPR), true negative rate (TNR), area under the ROC curve (AUC), positive predictive value (PPV), and negative predictive value (NPV).

In the 30-channel configuration, the features PLI, Phase-Locking Value (PLV), RP-I, and RP-II all achieved perfect classification rates (100%) in the training set, indicating excellent discriminative ability. Among these, PLI maintained robust performance in the test set, with a CR of 73.33%, TPR of 80.00%, and TNR of 66.67% (Table 3).

### Comparative Evaluation of Six Classifiers for Predicting rTMS Response in Depression Using 8- and 30-Channel EEG Features

In this study, six different classifiers—K-Nearest Neighbors (KNN), Linear Discriminant Analysis (LDA), Naive Bayes, Logistic Regression, Support Vector Machine (SVM), and eXtreme Gradient Boosting (XGBoost)—were employed to model the complete feature set, and their performances were evaluated under both 8-channel (8CH) and 30-channel (30CH) EEG configurations. The classification outcomes, including classification rate (CR), true positive rate (TPR), true negative rate (TNR), area under the curve (AUC), positive predictive value (PPV), and negative predictive value (NPV), are summarized in the table.

Under the 8CH configuration, LDA achieved the best performance on the training set, with a CR of 95.89%, TPR of 97.50%, and TNR of 94.29%. On the test set, LDA maintained solid generalization with a CR of 78.89%, TPR of 80.00%, and TNR of 77.78%. In contrast, XGBoost demonstrated a training CR of 84.11%, TPR of 90.00%, and TNR of 78.21%. However, after 1,000 random shuffles, XGBoost’s mean training CR dropped to 63.79% ± 4.61%, with TPR and TNR declining to 69.33% ± 6.53% and 58.25% ± 6.84%, respectively. On the test set, XGBoost achieved a CR of 72.78%, TPR of 90.00%, and TNR of 55.56%, indicating comparatively weaker generalization than LDA. The instability observed with XGBoost may be attributed to the limited training sample size (n = 75), which may be insufficient for adequately training a tree-based ensemble method such as XGBoost, known to require larger datasets to avoid overfitting and improve model robustness.

Under the 30CH configuration, LDA achieved a perfect classification performance on the training set, with 100% accuracy, sensitivity, and specificity, and demonstrated stable generalization on the test set with a CR of 73.33%, TPR of 80.00%, and TNR of 66.67%. SVM also performed well, achieving a training CR of 94.82% and a test CR of 83.89%, with TPR and TNR of 90.00% and 77.78%, respectively, and the highest AUC value of 86.67% among all classifiers tested.

The XGBoost classifier, under the 30CH configuration, initially achieved a training CR of 82.50%, with TPR of 87.14% and TNR of 77.86%. After 1,000 shuffles, its mean training CR was 69.57% ± 4.11%, with TPR and TNR dropping to 72.46% ± 5.43% and 66.68% ± 6.23%, respectively. On the test set, XGBoost yielded a CR of 73.33%, TPR of 80.00%, and TNR of 66.67%, with an AUC of 73.33%. Although XGBoost showed relatively good test performance, its lower stability during training again suggests that model performance may be hindered by the limited data size used in this study.

## Discussion

To the best of our knowledge, this is the first study to compare 8-channel and 30-channel EEG configurations for predicting treatment response to repetitive transcranial magnetic stimulation (rTMS) in patients with major depressive disorder (MDD). Using the 8-channel configuration, the model achieved a test classification accuracy of 78.89%, whereas the 30-channel configuration yielded a slightly lower accuracy of 73.33% on the test set. Models incorporating multiple features consistently outperformed those based on single features. Among the six classifiers evaluated, Linear Discriminant Analysis (LDA) demonstrated superior generalization and stability, particularly under conditions of limited training data.

### Superior Generalization of the 8-Channel Model

Previous studies have used high-density EEG to forecast rTMS outcomes ranged from 68.5% to 91.3%, with a pooled accuracy of 85.70% (95% CI: 77.45–94.83) (Bailey et al., 2018; Bailey et al., 2019; Corlier et al., 2019; Erguzel et al., 2015; Hasanzadeh et al., 2019). Our findings demonstrate that both the 8-channel and 30-channel EEG configurations yielded comparable predictive accuracy on the test set, achieving 78.89% and 73.33% respectively. This suggests that increasing the number of electrodes may enhance model fitting during training but does not necessarily improve generalization to unseen data. Another possible explanation is that relevant neural signatures predictive of treatment response may be spatially localized, and thus adequately captured by a carefully selected subset of electrodes(Wu et al., 2022; McCann et al., 2015). The comparable performance of the 8-channel model highlights the potential of targeted EEG acquisition strategies for clinical applications. Moreover, features such as phase synchronization (PLI and PLV) were consistently informative across both configurations, supporting prior evidence that altered functional connectivity is a key biomarker for rTMS treatment response in depression (Miljevic et al., 2023).

### Advantages of Multi-Feature over Single-Feature Model**s**

Previous rTMS trials, key predictive features have included absolute and relative power in frontal electrodes—particularly within the alpha and theta bands—as well as connectivity measures in the theta and gamma ranges. Additional relevant features comprise spectral entropy and cordance values across alpha, theta, delta, and gamma frequency bands(Watts et al., 2022). In our study some single EEG features—such as PLI and RP-I—showed relatively high classification performance, none exceeded 80% accuracy on the test set when used in isolation. This highlights the limitation of relying on individual features to predict treatment response. In contrast, our multi-feature models, constructed through a two-stage selection strategy (RFE-SVC followed by SBS with LDA classification), significantly improved predictive performance. The integration of diverse features—including spectral power, coherence, and phase-based connectivity metrics—likely captures complementary aspects of neural dynamics that contribute to treatment outcomes.

### LDA Outperforms XGBoost in Generalization and Stability with Limited EEG Data

In this study, Linear Discriminant Analysis (LDA) consistently outperformed XGBoost in terms of generalization and result stability when applied to a limited EEG dataset. With only 75 EEG data points available for training, LDA achieved a training classification rate (CR) of 95.89% and maintained solid test performance (CR = 78.89%), while XGBoost, despite an initially reasonable training CR of 84.11%, showed a significant drop in average performance after 1,000 shuffles (mean CR = 63.79% ± 4.61%). One possible explanation is that XGBoost, as a tree-based ensemble learning method, is highly data-dependent and prone to overfitting when the dataset is small or imbalanced(Zhang et al., 2025). Second, LDA’s linear and parametric nature allows it to converge reliably even in limited-sample scenarios, particularly when feature selection has already reduced dimensionality(Li et al., 2025). Third, LDA is less sensitive to hyperparameter tuning compared to XGBoost, which requires careful adjustment of multiple parameters (e.g., learning rate, tree depth) to achieve optimal performance—often difficult with small datasets(Dwyer et al., 2018). Overall, these findings suggest that inEEG-based predictive modeling, particularly when training data is limited, simpler linear models like LDA may offer better generalization and reliability than more complex machine learning algorithms such as XGBoost.

### Strength and implication

One notable strength of this study is its clinical applicability. The 8-channel configuration aligns with a commercially available, TFDA-approved EEG system, thereby enhancing the translational potential of our model. Moreover, the reduced electrode count offers practical benefits such as shorter setup time, increased patient comfort, and lower operational costs—factors that are critical for routine deployment in outpatient or psychiatric settings. Our two-stage feature selection method, combined with repeated shuffle validation, also enhances the reliability and stability of the model’s predictions. By demonstrating that a simplified EEG system can match the performance of a more complex setup, our findings pave the way for scalable and accessible diagnostic tools to support precision psychiatry. Furthermore, the study bridges a gap between AI modeling and regulatory-approved clinical tools, setting a precedent for future research aimed at integrating neurotechnology into evidence-based mental health care.

### Study Limitations

Despite promising results, several limitations should be acknowledged. First, the study’s relatively small sample size may limit the generalizability of the findings. While we employed 1,000 random shuffles to stabilize model estimates, larger multicenter datasets are needed to validate the robustness of the proposed approach. Second, our classification models were based primarily on LDA and SVC frameworks. While these algorithms provide interpretability and are well-suited for small samples(Cooney et al., 2020), future work should explore the potential of more advanced machine learning techniques such as random forests, gradient boosting, or deep learning architectures. Additionally, we did not examine the longitudinal dynamics of EEG features over the course of treatment, which could offer further insights into neuroplasticity associated with TMS. Addressing these limitations in future studies could help refine prediction models and broaden their utility in personalized treatment planning.

## Conclusions

This study highlights the potential of using pre-treatment EEG features to predict rTMS treatment response in patients with major depressive disorder. Using a two-stage AI model with feature selection and LDA classification, we found that an 8-electrode configuration achieved comparable—sometimes better—predictive accuracy than a 30-electrode setup on test data. This suggests that more electrodes do not necessarily yield better generalization, and simplified EEG systems may offer cost-effective, clinically viable solutions. Moreover, combining multiple nonlinear features significantly outperformed single-feature models, supporting the importance of multivariate neurophysiological markers. These findings provide a foundation for future work on AI-driven precision psychiatry.

## Data Availability

All data produced in the present study are available upon reasonable request to the authors

## Acknowledgement

We appreciate all the patients who have participated in this study. The authors also acknowledge the PET-MRI support from the Department of Radiology at Tri-Service General Hospital. And we also gratefully acknowledge the assistance of HippoScreen Neurotech Corp. for their support in data analysis.

## References

1. Bailey NW, Hoy KE, Rogasch NC, et al. (2018) Responders to rTMS for depression show increased fronto-midline theta and theta connectivity compared to non-responders. Brain Stimul 11: 190–203.

2. Bailey NW, Hoy KE, Rogasch NC, et al. (2019) Differentiating responders and non-responders to rTMS treatment for depression after one week using resting EEG connectivity measures. J Affect Disord 242: 68–79.

3. Brunoni AR, Chaimani A, Moffa AH, et al. (2017) Repetitive Transcranial Magnetic Stimulation for the Acute Treatment of Major Depressive Episodes: A Systematic Review With Network Meta-analysis. JAMA Psychiatry 74: 143–152.

4. Chung TH, Hanley K, Le YC, et al. (2023) A validation study of PHQ-9 suicide item with the Columbia Suicide Severity Rating Scale in outpatients with mood disorders at National Network of Depression Centers. J Affect Disord 320: 590–594.

5. Cooney C, Korik A, Folli R, et al. (2020) Evaluation of Hyperparameter Optimization in Machine and Deep Learning Methods for Decoding Imagined Speech EEG. Sensors (Basel) 20.

6. Corlier J, Wilson A, Hunter AM, et al. (2019) Changes in Functional Connectivity Predict Outcome of Repetitive Transcranial Magnetic Stimulation Treatment of Major Depressive Disorder. Cereb Cortex 29: 4958–4967.

7. Dwyer DB, Falkai P and Koutsouleris N. (2018) Machine Learning Approaches for Clinical Psychology and Psychiatry. Annu Rev Clin Psychol 14: 91–118.

8. Erguzel TT, Ozekes S, Gultekin S, et al. (2015) Neural Network Based Response Prediction of rTMS in Major Depressive Disorder Using QEEG Cordance. Psychiatry Investig 12: 61–65.

9. Fingelkurts AA and Fingelkurts AA. (2015) Altered structure of dynamic electroencephalogram oscillatory pattern in major depression. Biol Psychiatry 77: 1050–1060.

10. Garcia-Marin LM, Mulcahy A, Byrne EM, et al. (2023) Discontinuation of antidepressant treatment: a retrospective cohort study on more than 20,000 participants. Ann Gen Psychiatry 22: 49.

11. Gaynes BN, Rush AJ, Trivedi MH, et al. (2008) The STAR*D study: treating depression in the real world. Cleve Clin J Med 75: 57–66.

12. Hallett M. (2007) Transcranial magnetic stimulation: a primer. Neuron 55: 187–199.

13. Hasanzadeh F, Mohebbi M and Rostami R. (2019) Prediction of rTMS treatment response in major depressive disorder using machine learning techniques and nonlinear features of EEG signal. J Affect Disord 256: 132–142.

14. Kennedy SH, Lam RW, McIntyre RS, et al. (2016) Canadian Network for Mood and Anxiety Treatments (CANMAT) 2016 Clinical Guidelines for the Management of Adults with Major Depressive Disorder: Section 3. Pharmacological Treatments. Can J Psychiatry 61: 540–560.

15. Klooster D, Voetterl H, Baeken C, et al. (2024) Evaluating Robustness of Brain Stimulation Biomarkers for Depression: A Systematic Review of Magnetic Resonance Imaging and Electroencephalography Studies. Biol Psychiatry 95: 553–563.

16. Lefaucheur JP, Aleman A, Baeken C, et al. (2020) Evidence-based guidelines on the therapeutic use of repetitive transcranial magnetic stimulation (rTMS): An update (2014-2018). Clin Neurophysiol 131: 474–528.

17. Li Z, Nie F, Wang R, et al. (2025) A Revised Formation of Trace Ratio LDA for Small Sample Size Problem. IEEE Trans Neural Netw Learn Syst 36: 5803–5809.

18. Malhi GS and Mann JJ. (2018) Depression. Lancet 392: 2299–2312.

19. McCann MT, Thompson DE, Syed ZH, et al. (2015) Electrode subset selection methods for an EEG-based P300 brain-computer interface. Disabil Rehabil Assist Technol 10: 216–220.

20. Miljevic A, Bailey NW, Murphy OW, et al. (2023) Alterations in EEG functional connectivity in individuals with depression: A systematic review. J Affect Disord 328: 287–302.

21. Ricciardi C, Valente AS, Edmund K, et al. (2020) Linear discriminant analysis and principal component analysis to predict coronary artery disease. Health Informatics J 26: 2181–2192.

22. Saeys Y, Inza I and Larranaga P. (2007) A review of feature selection techniques in bioinformatics. Bioinformatics 23: 2507–2517.

23. Tang C, Gao T, Li Y, et al. (2022) EEG channel selection based on sequential backward floating search for motor imagery classification. Front Neurosci 16: 1045851.

24. Toussaint A, Husing P, Gumz A, et al. (2020) Sensitivity to change and minimal clinically important difference of the 7-item Generalized Anxiety Disorder Questionnaire (GAD-7). J Affect Disord 265: 395–401.

25. Watts D, Pulice RF, Reilly J, et al. (2022) Predicting treatment response using EEG in major depressive disorder: A machine-learning meta-analysis. Transl Psychiatry 12: 332.

26. Wu CT, Huang HC, Huang S, et al. (2021) Resting-State EEG Signal for Major Depressive Disorder Detection: A Systematic Validation on a Large and Diverse Dataset. Biosensors (Basel) 11.

27. Wu W, Ma L, Lian B, et al. (2022) Few-Electrode EEG from the Wearable Devices Using Domain Adaptation for Depression Detection. Biosensors (Basel) 12.

28. Yen YC, Chiu NY, Hwang TJ, et al. (2022) A Multi-Center Study for the Development of the Taiwan Cognition Questionnaire (TCQ) in Major Depressive Disorder. J Pers Med 12.

29. Zhang L, Jian L, Long Y, et al. (2025) Machine learning approaches for classifying major depressive disorder using biological and neuropsychological markers: A meta-analysis. Neurosci Biobehav Rev 174: 106201.

